# AUTOMATED VERSUS MANUAL REANALYSIS IN RARE DISEASE GENOMICS

**DOI:** 10.64898/2026.05.16.26352295

**Authors:** Daniel Kaschta, Vincent Arriens, Simon Müller, Caroline Utermann-Thüsing, Genom-RD, Inga Vater, Almuth Caliebe, Inga Nagel, Malte Spielmann

## Abstract

**Purpose:** Periodic reanalysis of genome sequencing data can yield additional diagnoses as knowledge evolves, yet manual reanalysis is labour-intensive. We compared automated and manual reanalysis approaches in rare disease genomics.

**Methods:** We reanalyzed 377 rare disease cases: 158 with pathogenic or likely pathogenic (P/LP) findings, 49 with variants of uncertain significance (VUS) findings, and 170 had no findings. Manual reanalysis used standard diagnostic workflow for all cases without prior P/LP diagnoses (219 cases). An automated pipeline using Talos was benchmarked on the 158 P/LP cases before application to the 219-case reanalysis cohort. The mean reanalysis interval was 660 days.

**Results:** Manual reanalysis identified three additional P/LP cases and two newly classified as VUS, increasing P/LP cases from 158 (41.9%) to 161 (42.7%). Talos recovered all three P/LP findings but only identified one of the two new VUS findings. Benchmarking showed 80.0% singleton concordance and 75.2% (82.8% proband-only) trio concordance, with an output of approximately three variants per case.

**Conclusion:** Reanalysis at 1.8 years yields modest but clinically meaningful gain. Automated reanalysis closely approximates manual performance while reducing hands-on effort, supporting scalable reanalysis in routine genomic care.

## 1. Introduction

In recent years, short-read genome sequencing (GS) has become a first-line diagnostic approach for individuals with rare genetic disorders in many diagnostic settings. However, its initial diagnostic yield remains limited to approximately 45%, leaving a substantial proportion of cases without reported candidate variants [1]. Compared with exome sequencing, GS can overcome some technical limitations through more uniform coverage and improved detection of short tandem repeats (STRs), noncoding, copy-number variants (CNVs), and mitochondrial variation [2–5]. This results in comprehensive genomic data that can be stored and repeatedly reanalyzed as knowledge and methods improve.

The continuous expansion of gene-disease associations, updates to variant classification frameworks, and improvements in analytics and annotation resources provide a strong rationale for the reanalysis of existing genomic data [6]. Reanalysis is now recommended as part of routine rare-disease diagnostics and can deliver clinically relevant diagnoses years after the original analysis [7]. Meta-analytic evidence suggests that reanalysis yields additional diagnoses over time, with an overall diagnostic yield of 10% (95% CI 6–13%) across studies, and a higher yield of 13% for reanalysis con ducted ≥24 months after original analysis [8]. However, this estimate is based on highly heterogeneous studies that differ in sequencing modality, reanalysis intervals, initial cohort diagnostic yield, data reuse strategy, and degree of human oversight, limiting its applicability to routine clinical practice [8].

Despite its potential, the implementation of reanalysis remains significantly constrained by the substantial hands-on time and the personnel it demands. Complex clinical and laboratory processes, limited reimbursement, workforce shortages, and, in some settings, inadequate data infrastructure or additional fees further exacerbate these challenges [9]. As a result, access to reanalysis remains low and inequitable, often depending on particularly motivated clinicians, laboratories, or patients [9, 10].

Traditionally, reanalysis has relied on expert-driven manual interpretation, which is effective but labor-intensive and difficult to scale as sequencing volumes increase [9]. Automated reanalysis frameworks have emerged to address this bottleneck by systematically reannotating and reprioritizing variants as knowledge evolves, potentially enabling more frequent and timely reinterpretation [11]. However, automated reanalysis models demand explicit trade-offs between sensitivity, specificity, reanalysis frequency, and degree of automation [12, 13]. Their implementation must also balance clinical impact against additional workload and financial reimbursement, and depends on adequate bioinformatics and health-informatics infrastructure to support increased data volumes [9, 14]. In addition, automated reanalysis raises ethical, legal, and logistical questions, including consent, patient recontact, and responsibility for acting on newly identified findings [9, 15, 16].

Direct comparisons between manual clinical reanalysis and automated, phenotype aware pipelines remain scarce, particularly in real-world hospital settings using archived diagnostic genome data. Whether automated approaches can reproduce clinically accepted expert reclassifications, where they fall short, and what workload they introduce in practice are poorly understood. Large-scale, non-selective cohort evaluations are needed to assess performance, implementation burden, and to guide laboratory practice, clinical adoption, and funding policy.

Here, we compared manual reanalysis in a clinical diagnostic setting with automated reanalysis using the open-source tool Talos [13] in a well-characterized rare-disease genome sequencing cohort. Using archived diagnostic data, we assessed concordance with expert review, incremental diagnostic yield, and the strengths and limitations of automation, providing evidence for the integration of scalable reanalysis strategies into routine clinical genomics.

## 2. Methods

### 2.1 Study Cohort and Reanalysis Design

In this comparative GS analysis, we revisited cases of individuals with suspected rare diseases originally recruited through routine clinical care at the University Medical Center Schleswig-Holstein (UKSH) [17] as part of an extended rare-disease genome sequencing initiative. All participants provided written informed consent, allowing genome sequencing and subsequent reanalysis of genomic data.

For this reanalysis study, we included 377 index cases, comprising 305 trio cases and 72 singleton cases. Cases were stratified according to the outcome of the original diagnostic analysis. Within the singleton cohort, 29 cases harbored results based on pathogenic or likely pathogenic (P/LP) variants, 16 were cases with VUS (variant of uncertain significance) findings, and 27 had no findings. Within the trio cohort, 129 cases harbored results based on P/LP variants, 33 were cases with VUS findings, and 143 had no findings.

Cases with confirmed results based on P/LP variants and cases with VUS findings from both singleton and trio cohorts were used as a benchmarking set for evaluating the performance of automated reanalysis using Talos (version 8.2.0). For reanalysis outcome assessment and variant reassessment, cases without a prior P/LP diagnosis (cases based on no-finding and VUS findings) constituted the primary reanalysis cohort for manual and automated strategies. The mean time interval between the initial genome analysis and reanalysis was 660 days.

### 2.2 Genome Sequencing, Primary Processing, and Quality Control

All reanalyses were performed using archived genome sequencing data originating from the initial analysis. GS was performed on DNA (deoxyribonucleic acid) extracted from peripheral blood samples. Libraries were prepared using PCR-free protocols and sequenced on Illumina NovaSeq instruments with paired-end 150-bp reads, targeting genome-wide coverage consistent with diagnostic whole-genome sequencing. Sequence reads were aligned to the human reference genome (GRCh38) and processed using the Illumina DRAGEN™ germline pipeline v3.7.5 [18]. Variant calling included the detection of single-nucleotide variants (SNVs), small insertions and deletions, copy-number variants (CNVs) [19], and short tandem repeats (STRs) [20] using the corresponding DRAGEN modules. Quality control filters were applied to ensure sufficient read depth, variant allele balance, overall call confidence, and a minimum anticipated genome wide coverage (mean mapped read depth) of 29-fold per case; the actual documented mean coverage was 38× throughout the cohort.

### 2.3 Variant Annotation and Interpretation

Variant annotation and interpretation followed the same framework as the original cohort analysis. Variants were annotated using current versions of population frequency databases, clinical variant repositories, and gene-disease resources, including gnomAD [21], ClinVar [22], OMIM [23], DECIPHER [24], and PubMed [25]. Phenotype–genotype correlation was guided by Human Phenotype Ontology (HPO) [26] terms. Variant classification followed the guidelines of the American College of Medical Genetics and Genomics (ACMG) [27], the Association for Molecular Pathology (AMP) [28], and the Association for Clinical Genomic Science (ACGS) [29]. All variant descriptions were validated for accuracy and conformance to HGVS nomenclature using VariantValidator software [30]. Diagnostic yield was categorized into three classes:

1. P/LP: A pathogenic or likely pathogenic variant explaining the patient’s phenotype and consistent with the inheritance pattern was identified.
2. VUS: A variant of uncertain significance explaining the patient’s phenotype and consistent with the inheritance pattern was identified, with strong supporting evidence (“hot VUS”).
3. No findings: no clinically relevant diagnostic variants were identified.

A case was reported to the participants if the newly identified variants were classified as clinically relevant [P/LP or VUS] according to ACMG/AMP criteria and provided a coherent molecular explanation for the patient’s phenotype. For 2-hit cases carrying one P/LP variant and a VUS, case-level classification was determined by clinical assessment and recorded in the case metadata.was 38× throughout the cohort.

### 2.4 Manual Reanalysis of Previously No-Finding and Cases with VUS Findings

Manual reanalysis was performed using Illumina Emedgene [31] for all cases lacking a prior P/LP diagnosis (i.e., cases with VUS findings and no-finding cases). The original raw FASTQ files were reanalyzed, including repeat alignment and variant calling with an updated Illumina DRAGEN germline pipeline version (v4.2.4 for small-variant calling; v4.2 for survival motor neuron (SMN) copy-number analysis, STR calling, and SV calling)[18], rather than reuse of previously generated variant call files. No additional resequencing was performed.

Updated variant calls were annotated and interpreted in the Emedgene platform using current annotation resources, updated population allele frequencies, curated clinical information for refined phenotype–genotype matching, and the latest literature, with particular attention to newly established or reclassified gene–disease associations.

The average time required for full manual case reanalysis using Emedgene was also recorded.throughout the cohort.

### 2.5 Manual Reassessment of Previously Reported VUS

Previously reported VUS variants were systematically reassessed to determine whether newly available evidence supported reclassification. Evaluation criteria included newly available functional or clinical data, revised population allele frequency information, updated inheritance or segregation evidence, and the identification of additional variants contributing to a plausible molecular diagnosis. Reassessment was performed in accordance with updated ACMG/AMP variant classification criteria; however, final reevaluation was restricted to variants for which the accumulated evidence potentially supported reclassification to P/LP.

### 2.6 Automated Reanalysis Using Talos

Automated reanalysis was performed using Talos [13], an open-source phenotype aware platform for iterative rare-disease genome interpretation. Talos applies rule based variant prioritization using curated gene–disease resources (including PanelApp [32] and ClinVar [22]), inheritance-aware filtering, consequence-based prioritization, and configurable ACMG-aligned logic. The pipeline reannotates variants using current reference datasets and identifies variants that have become reportable based on updated clinical or gene–disease evidence since the previous analysis.

Talos was executed largely according to the developers’ recommended workflow, using archived per-case variant call format (VCF) files (compound index and relatives data), pedigree information where available, and optional HPO terms.

To enable the reuse of archived VCFs generated during the initial diagnostic analysis, the Talos workflow was customized to accept VCFs that had already been annotated by the Illumina DRAGEN pipeline. This adaptation allowed the integration of previously generated DRAGEN annotations into the Talos prioritization framework without recalling variants.

Talos was run without manual curation. The output consisted of structured candidate variant reports generated per case, which were directly compared with the results from manual reanalysis. According to the documentation, this modified version:

- preserves structural-variant coordinates during merging,
- adds gene annotation for structural variants using GFF3 overlap detection,
- handles CNVs appropriately by converting (CNV)-tag to (DEL) or (DUP), and
- decomposes multiallelic structural variants for compatibility with downstream tools.

### 2.7 Concordance Analysis and Error Types

Concordance was defined as the automated identification of variants that were newly detected or reclassified through manual reanalysis. Variants identified manually but not returned by Talos were considered missed by automated reanalysis. To investigate the causes of missed P/LP findings, discrepancies were grouped into five error categories:

1. Out-of-scope variant classes. Variants were considered out of scope when they fell outside the current capabilities of the Talos workflow. This includes, for example, mitochondrial variants and short tandem repeat (STR) expansions, which are not comprehensively handled by the present pipeline implementation.
2. Phenotype annotation gap. Some variants were not prioritized because the phenotypic similarity between the participant’s HPO profile and the gene associated phenotype was insufficient for prioritization. These cases likely reflect incomplete, imprecise, or non-overlapping phenotype annotations rather than an absence of clinical relevance.
3. Pipeline processing or conversion errors. Some variants were excluded because they were not successfully processed during file conversion or downstream pipeline steps. Consequently, these variants were not represented correctly in the Talos input or could not be prioritized appropriately.
4. Large CNVs. Complex copy-number variants spanning multiple genes were not consistently prioritized by Talos. These events represented a distinct technical limitation of the workflow, particularly when variant interpretation depended on a broader genomic context rather than single-gene assignment.
5. MOI filtering in trio analysis. In some trio cases, variants were excluded because inheritance and mode-of-inheritance information was interpreted too stringently by the automated pipeline, for example, by not accounting for reduced penetrance or atypical segregation. In these cases, rerunning the analysis without parental information restored variant prioritization, indicating that the filtering logic, rather than the variant itself, accounted for the missed call.

### 2.8 Benchmarking of Automated Reanalysis

Talos performance was independently benchmarked using a positive control set derived from the original cohort, comprising 129 trio cases and 29 singleton cases with known P/LP variants and 34 trio cases and 16 singleton cases with VUS variants. For each case and its associated reported variants in the benchmarking set, the fate of each individual variant, whether retained or excluded, was tracked independently. The detection rate was defined as the proportion of cases in which Talos successfully prioritized the known P/LP diagnostic variant and was calculated either in the merged cohort or separately for trios and singletons.

### 2.9 Language Editing and AI-Assisted Copy Editing

ChatGPT (OpenAI) was used exclusively for copyediting to improve readability and language [33]. The authors reviewed and refined all content and assume full responsibility for the final manuscript.

## 3. Results

### 3.1 Cohort

Between January 2022 and April 2023, 416 index cases comprising 1,011 individuals, including relatives, were recruited, sequenced and analyzed (Supplementary Table S1). Following verification of data availability, 377 cases comprising 920 genomes were retained for reanalysis (Fig. 1A). These included 305 trios or other multi-member families and 72 singleton cases. The cohort at sequencing was predominantly male (66.3%), and most patients were younger than 20 years of age (84.4%; Fig. 1B). The interval between the initial analysis and reanalysis ranged from 208 to 1,208 days, with a mean of 660 days (approximately 1.8 years; Standard Diviation: 257 days; Fig. 1C) (Supplementary Table S1).

**Fig. 1.**
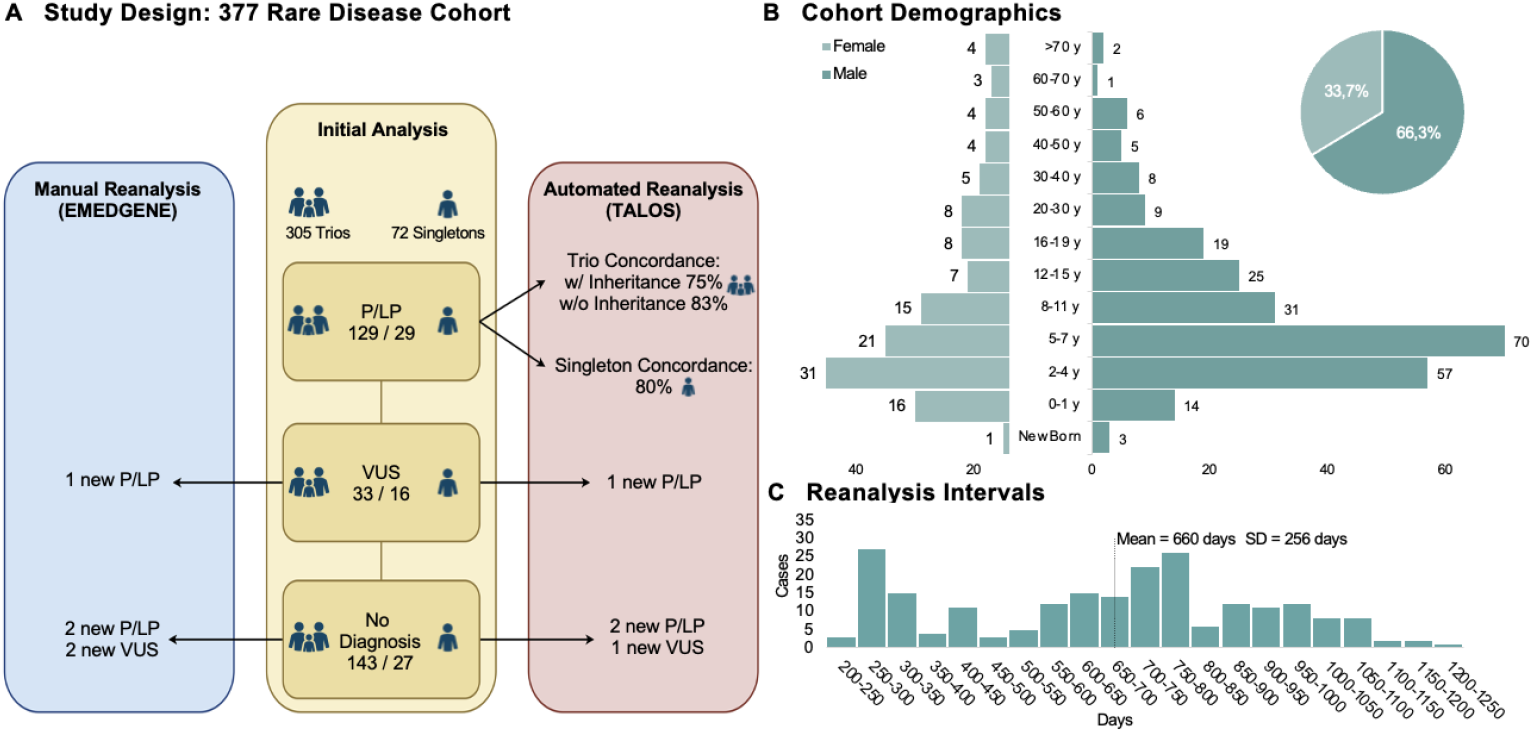
Study design and cohort overview for reanalysis. **(A)** We reanalyzed 920 individuals from 377 index cases (305 trios/multi-member, 72 singletons) with varied diagnostic outcomes using manual and automated workflows. Manual reanalysis identified one new pathogenic/likely pathogenic (P/LP) diagnosis in a case with VUS findings (also detected automatically), two further P/LP variants, and two VUS (one missed by the automated workflow) in cases without a prior diagnosis. **(B)** Sex and age distribution of 377 patients with available demographic data (250 males, 127 females). **(C)** The mean time from the initial analysis to reanalysis was 660 days.

At the time of the initial analysis, 158 cases (41.9%) had a P/LP variant, 49 cases (13.0%) were cases with VUS findings, and 170 cases (45.1%) had no reported molecular diagnosis. Reanalysis was performed using two complementary approaches. Manual reanalysis, conducted between October 2024 and December 2025 using Illumina Emedgene, was applied to all cases lacking a prior P/LP diagnosis (i.e., the 49 cases with VUS findings and 170 no-finding cases). An automated reanalysis pipeline based on an adapted version of Talos was applied to the entire cohort in October 2025, enabling a direct comparison with the manual approach.

Cases with prior P/LP results or cases with VUS results served as a benchmark set to evaluate the performance of the automated pipeline.

### 3.2 Benchmarking of Automated Reanalysis: Singleton Cases

Of the 45 singleton cases in the benchmarking set, 29 cases carried 35 P/LP variants underlying the initial diagnosis and 16 cases with VUS findings carried 21 VUS variants (Fig. 2). The fate of each individual variant, retained as reported as P/LP or VUS or potentially reclassified during reanalysis, was tracked independently using ACMG criteria.

**Fig. 2.**
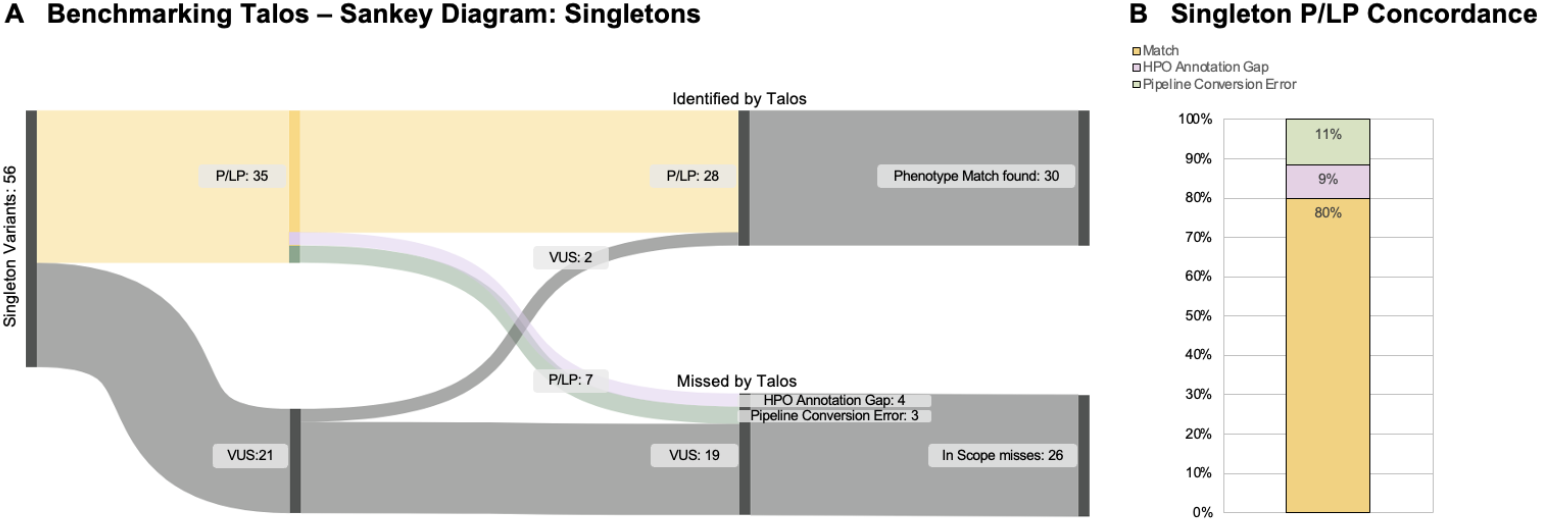
Benchmarking Talos with singleton cases. **(A)** Sankey diagram showing Talos prioritization performance across 45 singleton benchmarking cases. The input variants (left) comprised 35 pathogenic/likely pathogenic (P/LP) and 21 variants of uncertain significance (VUS). Of these, Talos successfully prioritized 28 P/LP and 2 VUS, with 30 phenotype matches; 7 P/LP and 19 VUS were missed (26 in-scope misses total). **(B)** Singleton concordance of missed P/LP variants broken down by failure mode: No Phenotype Match (11.4%), Pipeline Conversion (9.6%) and successfully matched (80.0%).

Talos prioritized 28 out of 35 P/LP variants (80.0% concordance), whereas only two out of 21 VUS were captured, which is consistent with the pipeline’s design preference for high-evidence variants. All prioritized variants were concordant with the recorded HPO-based phenotype annotations.

The seven missed P/LP variants were attributable to HPO annotation gaps (11.4%), in which the gene–phenotype association was not captured with sufficient specificity in the available ontology resources, and to pipeline conversion errors (8.6%) that prevented correct variant mapping within the prioritization framework (Supplementary Table S2).

### 3.3 Benchmarking of Automated Reanalysis: Trio Cases

Of the 162 trio benchmarking cases, 129 cases were reported with 145 P/LP variants and 33 cases with 43 VUS variants (Fig. 3). Talos prioritized 109 P/LP variants, for responding to an overall P/LP concordance of 75.2%. Six VUS variants were captured. All prioritized variants were concordant with recorded phenotype annotations. Failure modes for missed P/LP variants included four out-of-scope variant classes (2.8%), three large copy-number variants not processed by the pipeline (2.1%), nine pipeline conversion errors (6.2%), and nine HPO annotation gaps (6.2%) (Supplementary Table S2). Four P/LP variants were outside the scope of Talos analysis, comprised of two short tandem repeats, one mitochondrial variant, and one mosaic variant not supported by the pipeline.

**Fig. 3.**
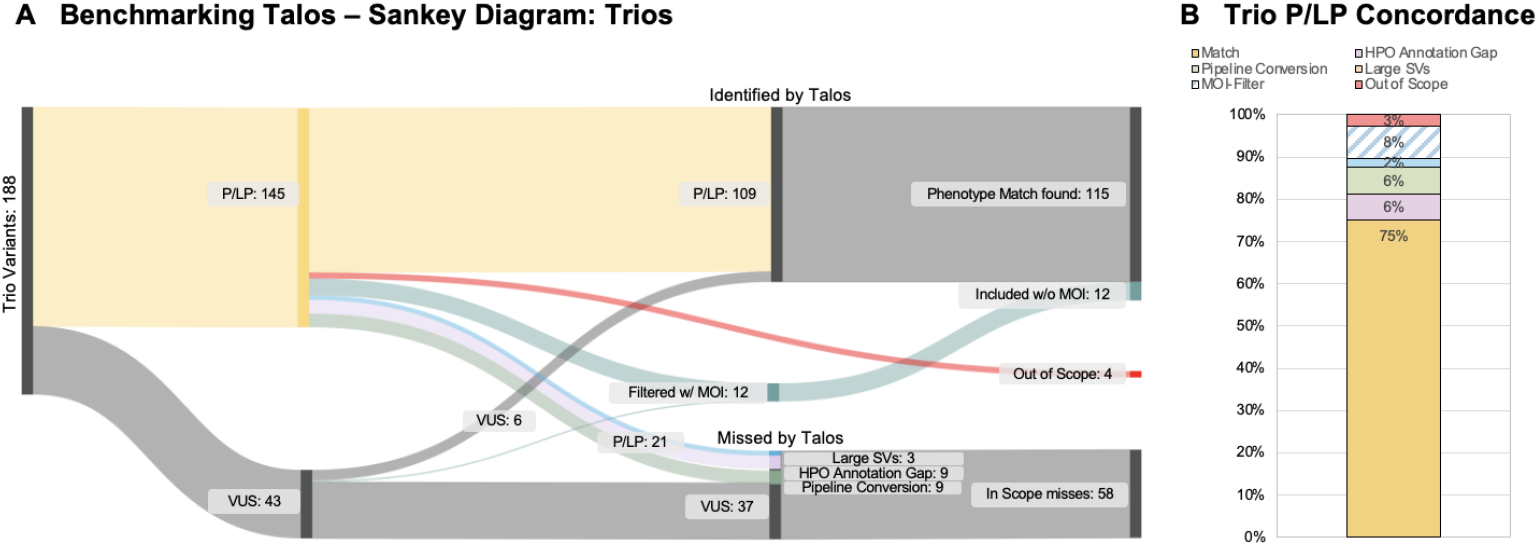
Benchmarking Talos with trio cases. **(A)** Sankey diagram showing Talos prioritization performance across 162 trio benchmarking cases. Input variants (left) comprised 145 pathogenic/likely pathogenic (P/LP) and 43 variants of uncertain significance (VUS). The pipeline identified 109 P/LP and six VUS, with 115 phenotype matches; 11 additional P/LP variants were recovered via proband only analysis after trio mode of inheritance (MOI) filtering. Four variants were out of scope. The “missed by Talos” variants comprised 21 P/LP and 37 VUS (58 in-scope misses total). **(B)** The right panel breaks down the missed P/LP variants by failure mode: successfully matched (75.2%), MOI filtered (7.6%), No Phenotype Match (6.2%), Pipeline Conversion (6.2%), large copy-number variant (CNV) not detected (2.1%), and out-of-scope variant class (2.8%). Percentages shown are rounded to the nearest integer.

An additional 11 P/LP variants (7.6%) were not prioritized when parental genotype data were included because Talos applied strict inheritance-based filtering, which excluded variants inconsistent with conventional dominant or recessive transmission models.

Notably, all 11 variants were successfully recovered when Talos was applied in the proband-only mode, increasing concordance to 82.8% and indicating a systematic limitation in the handling of reduced penetrance or atypical inheritance patterns under trio-mode analysis.

### 3.4 Reanalysis Diagnostic Yield: Manual Reanalysis

A manual reanalysis of the 49 cases with VUS findings and 170 no-finding cases identified five cases with newly reported findings, summarized in Table 1. Four of these cases had previously had no findings; in one case, a variant was newly classified as likely pathogenic based on recent literature, while the previously reported VUS remained unchanged. In case 92 (singleton), a pathogenic copy-number variant affecting *KANSL1* was not reported in the original analysis. Reanalysis with an updated tertiary variant-interpretation platform (Illumina Emedgene) prioritized the variant and supported its pathogenic classification.

**Table 1:** Variants identified during the rare-disease reanalysis. The table lists newly reported variants identified during manual reanalysis, including the case type, affected gene, HGVS nomenclature, case-level classification, status in the initial analysis, the main driver of re-identification, and whether the variant was also prioritized by Talos. Pathogenic or likely pathogenic **(P/LP);** Variant of uncertain significance (VUS). Case 228 carried two compound-heterozygous *COQ4* variants contributing to a single case-level diagnosis.

| Case | Type | Gene | Variant nomenclature (HGVS) | Class. | Initial analysis | Identified by | Talos |
| --- | --- | --- | --- | --- | --- | --- | --- |
| 92 | Singleton | KANSL1 | NC_000017.11:<br>g.46060362_46135409del<br>(75.05 kb) | P/LP | Unreported | Updated tertiary<br>analysis software | Match |
| 228 | Singleton | COQ4 | NC_000009.12:<br>g.128333549C>A<br>NM_016035.5:<br>c.702C>A<br>(p.Asn234Lys) | P/LP* | Unreported | Updated OMIM | Match |
| 228 | Singleton | COQ4 | NC_000009.12:<br>g.128323151A>C<br>NM_016035.5:<br>c.202+4A>C | P/LP | Unreported | Updated OMIM | Match |
| 101 | Trio | ZMYM3 | NC_000023.11:<br>g.71249681T>C<br>NM_201599.3:<br>c.1252-2A>G<br>(*de novo*) | VUS | Unreported | Updated OMIM /<br>GeneMatcher | Match |
| 354 | Trio | MYCBP2 | NC_000013.11:<br>g.77211220C>G<br>NM_015057.5:<br>c.3363G>C<br>(p.Glu1121Asp)<br>(*de novo*) | VUS | Unreported | New literature | No match |
| 39 | Trio | XPO1 | NC_000002.12:<br>g.61494067dup<br>NM_003400.4:<br>c.1072dupT<br>(p.Ser358PhefsTer2)<br>(*de novo*) | P/LP | Alternative VUS<br>reported | New literature | Match |

In cases 228 and 39, updated gene-disease association evidence from the current literature and databases supported the reclassification as P/LP under the ACMG/AMP criteria. Case 228 (singleton) carried two compound heterozygous variants in *COQ4* (case clinically classified as P/LP; one variant classified as VUS); both variants contributed to the P/LP case-level diagnosis. Case 39 (trio) harbored a *de novo XPO1* variant classified as LP, notably a different variant from the one originally reported as a VUS for this case. In cases 101 and 354 (both trios), newly available gene–disease association evidence supported clinical relevance; however, the evidence was insufficient for P/LP reclassification, and both variants were retained as clinically reportable VUS. The finding in case 101 (*ZMYM3, de novo*) was additionally supported by a GeneMatcher collaboration (manuscript in preparation) [34, 35].

Across the reanalysis cohort, manual review increased the number of cases with a reported P/LP finding from 158 to 161 (+0.8 percentage points), yielding a total diagnostic rate of 42.7%. The number of cases with VUS findings increased from 49 to 50, reflecting the addition of two newly identified VUS (cases 101 and 354), offset by the upgrade of patient 39 from VUS case to P/LP case based on the newly identified variant. Consequently, the number of cases without any reported clinically relevant findings decreased from 170 to 166 (-1.0%). For the 219 reanalysis cases (49 cases with VUS findings + 170 no-finding cases), we measured the mean time required for full manual case reanalysis. The mean reanalysis time was 81 minutes per case, not including report writing or the preparation of evidence for strong candidate variants for independent secondary review.

### 3.5 Reanalysis Diagnostic Yield: Automated Reanalysis

Automated reanalysis using the Talos pipeline also identified the three new P/LP cases, matching the manual P/LP gain in full, and prioritized the newly identified VUS in case 101(Table 1). The *de novo MYCBP2* variant in case 354, which had been identified during manual reanalysis, was not returned by the automated workflow, consistent with the pipeline’s limited prioritization of variants in genes lacking sufficient evidence in current curated databases. Automated reanalysis increased the number of cases with a P/LP finding from 158 to 161 (+0.8 percentage points; 41.9% total diagnostic yield), which is identical to the manual result. The number of cases based on results with VUS findings remained at 49, with one previously no-finding case (case 101) newly prioritized and offset by the reclassification of case 39 based on the newly identified variant. The number of cases without any prioritized finding decreased from 170 to 167 (-0.8 percentage points), one case fewer than the manual reduction of four no finding cases. On average, Talos returned three variants per case for singleton cases and trio cases (with parental information).

### 3.6 Comparison

Overall, both reanalysis strategies yielded similar gains in P/LP diagnoses. Manual reanalysis was more sensitive for the identification of clinically relevant VUS in emerging gene–disease associations, detecting one additional reportable VUS (case 354) that fell below the automated prioritization threshold. These results are summarized in Fig. 4, which illustrates the distribution of cases across outcome categories (P/LP, cases with VUS findings, and no diagnosis) at initial analysis, after manual reanalysis, and after automated reanalysis.

**Fig. 4.**
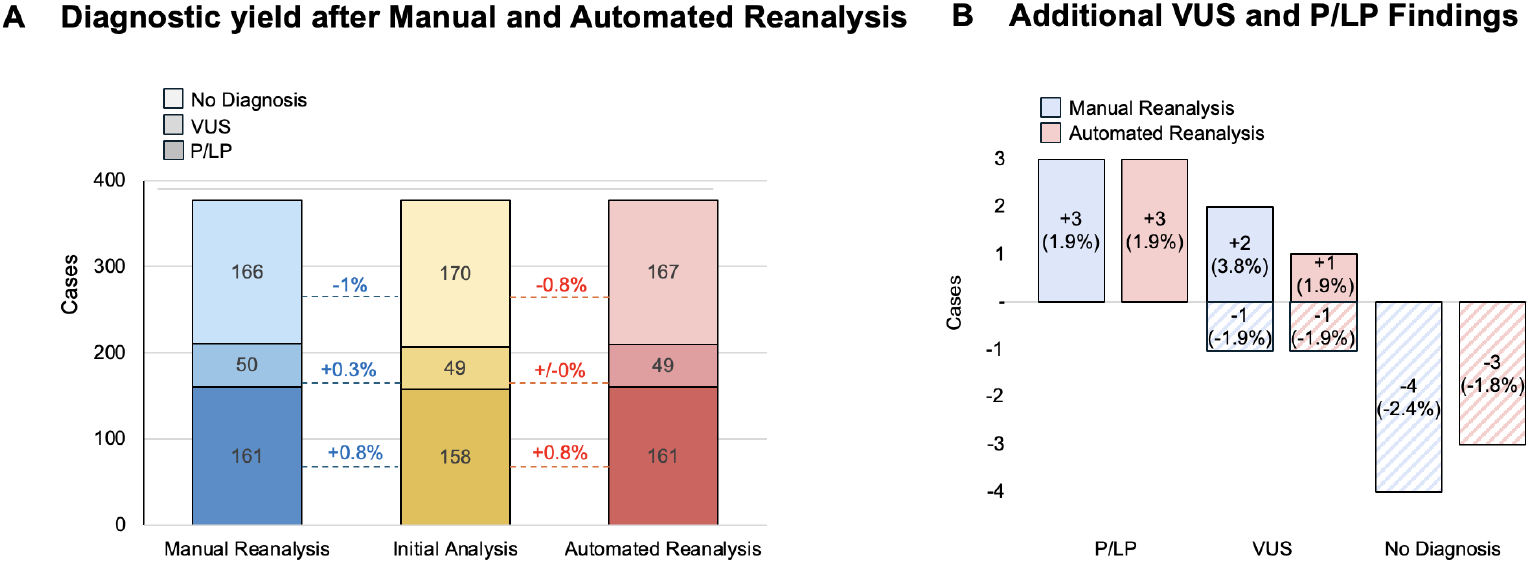
Diagnostic yield after manual and automated reanalysis. **(A)** Stacked bar charts showing case distribution across three categories (pathogenic/likely pathogenic [P/LP], cases with variants of uncertain significance [VUS] findings, and no molecular diagnosis) at initial analysis, manual reanalysis, and automated reanalysis. Bars display absolute counts; connecting lines between time points illustrate net category transitions and incremental diagnostic gain. Both workflows achieved identical P/LP gains (+3 cases; 41.9% to 42.7%). Manual reanalysis identified one additional case with VUS findings (49 vs. 50), reflecting greater sensitivity for emerging gene–disease associations. **(B)** Comparison of changes in case classification after manual and automated reanalysis. Bars above zero represent newly assigned findings, whereas hatched bars below zero represent cases removed from the respective category following reclassification. Both approaches yielded 3 additional P/LP findings. Manual reanalysis produced a larger increase in cases with VUS findings than automated reanalysis (two vs. one cases), whereas both approaches reclassified one case from the VUS category. Values within the bars denote absolute case number changes per category.

## 4. Discussion

Reanalysis of whole-genome sequencing data from 377 rare disease cases yielded incremental but clinically relevant improvements in diagnostic resolution. After a mean interval of 660 days (approximately 1.8 years), manual reanalysis identified three additional cases with P/LP findings, increasing the diagnostic yield from 41.9% to 42.7%, and reassigned two previously undiagnosed cases to the VUS category. The automated Talos-based workflow recapitulated all three P/LP findings and detected one additional VUS case, capturing all but one clinically relevant variant identified by manual review. These outcomes demonstrate that automated reanalysis closely approximates the diagnostic performance of expert-driven interpretation while substantially reducing interpretive burden. Automated triage to a median of approximately three candidate variants per case further supports its scalability for routine genomic reanalysis.

### 4.1 Diagnostic Yield in the Context of Published Reanalysis Studies

The absolute diagnostic gain was less than 1% for P/LP variants, markedly lower than the 5-20% reported in previous reanalysis studies [6–8]. This difference is largely attributable to the cohort characteristics and baseline performance. Prior studies typically include heterogeneous datasets (exome and genome), longer reanalysis intervals, and variable initial analysis quality, whereas our prospectively ascertained cohort underwent uniform short-read genome sequencing with a high initial diagnostic yield (41.9%) under standard-of-care conditions. This high baseline inherently limits the potential for further diagnostic uplift and likely reflects the current real-world ceiling of reanalysis in well-characterized clinical cohorts. Furthermore, no diagnostic gain was attributable to the correction of human interpretive errors from the initial analysis, compared to other studies. All newly identified P/LP findings and clinically relevant VUS arose exclusively from updated variant-calling and interpretation pipelines, revised gene–disease validity evidence, or newly published literature [36] and Gene eMatcher collaborations [34, 35]. This finding underscores that, in settings with a rigorous initial analysis, reanalysis of older sequencing data is driven less by quality improvement and more by the ongoing integration of new knowledge.

Of note, no previously reported VUS were reclassified as P/LP upon reanalysis. Although VUS upgrading is described as a key rationale for periodic reanalysis [37], it depends on the accumulation of robust functional, segregation, or population-level evidence. The absence of reclassification after approximately two years likely reflects the interval being insufficient for meaningful evidence accumulation. Published clinical genetics guidelines, including the 2024 ACGS Best Practice Guidelines for Variant Classification in Rare Disease, have provided clarified evidentiary thresholds for criteria such as PP4 (population frequency), which may contribute to more conservative classification strategies in reanalysis scenarios [29]. Taken together, the short interval and the changing evidence standards in published guidance likely account for the absence of VUS reclassification in this cohort.

### 4.2 Automated Reanalysis: Strengths and Limitations

Benchmarking of the Talos pipeline against known P/LP variants showed concordance rates of 80.0% for singletons and 75.2% for trios, increasing to 82.8% in proband-only mode. This is consistent with the original Talos study, which reported concordance rates of 69–81% across two test cohorts [13]. Discordant cases reflected multiple limitations rather than a single source of error.

First, technical constraints contributed to missed variants, including those outside the pipeline scope (e.g., mitochondrial variants and STR expansions) and reduced prioritization of large, multi-gene CNVs. In addition, conversion and integration issues with archived VCFs prevented 12 variants from entering the prioritization workflow, highlighting constraints related to technical scope and data compatibility. A more Talos-tailored VCF processing pipeline could reduce data compatibility constraints by ensuring more reliable variant ingestion and enabling more thorough identification and evaluation of candidate variants. Second, 13 missed variants were attributable to limitations of phenotype-driven prioritization, reflecting incomplete or nonspecific HPO annotation and gaps in gene–phenotype knowledge. Third, lower concordance in trios indicates a limitation of inheritance-based filtering. Rigid mode-of-inheritance rules led to the exclusion of 11 P/LP variants, particularly in the context of reduced penetrance or atypical inheritance. The higher concordance in proband-only mode suggests that trio-based filtering may introduce false negatives and should be applied more flexibly, with provision for secondary expert review. The single missed case involved a *de novo* variant in *MYCBP2*, identified manually but not prioritized due to insufficient phenotype–genotype evidence, illustrating a general limitation of rule-based frameworks in emerging disease genes. Regular updates of gene–disease resources and periodic review of borderline candidates are therefore essential.

Despite these limitations, automated reanalysis offers a major practical advantage through rapid turnaround and minimal hands-on time, substantially reducing expert workload and likely associated costs. In contrast to manual reanalysis, which required a mean of 81 minutes per case, the automated approach reduced review burden markedly by limiting assessment to an average of only three candidate variants per case. Importantly, the automated workflow recovered all newly identified P/LP findings without manual input, indicating that high-confidence diagnostic variants are reliably captured.

### 4.3 Implications for Clinical Practice

Although periodic reanalysis provides measurable diagnostic benefit, the absolute gain in our cohort remains modest and is often not reflected in current reimbursement structures [38], creating a gap between guideline recommendations and clinical feasibility. Automated triage offers a practical solution by focusing expert review on a small set of high-confidence candidates, thereby reducing workload while preserving clinical oversight. This tiered model supports scalability with increasing sequencing volumes and may be adjusted to allow broader review where capacity permits, potentially improving the diagnostic yield.

In parallel with economic optimization, broader adoption faces unresolved challenges around patient consent for iterative reanalysis, mechanisms for patient recontact, and institutional accountability when automated workflows surface newly reportable variants [9, 15, 16]. This study did not assess cost-effectiveness or address these ethical, legal, and logistical dimensions. Prospective economic evaluations and dedicated framework development remain prerequisites for the routine clinical integration of automated reanalysis.

## 5. Conclusion

Genomic reanalysis after a mean interval of 660 days (approximately 1.8 years) yields a clinically meaningful, if incremental, improvement in diagnostic resolution in a high-yield rare-disease cohort. The primary drivers of new diagnoses are advances in variant-calling and gene–disease association rather than retrospective correction of interpretive errors. This highlights the importance of infrastructure for continuous knowledge integration rather than one-time reanalysis events. Automated reanalysis using Talos closely approximates manual outcomes for high-confidence P/LP findings while concentrating expert effort on a small, prioritized candidate set. This supports automated triage as a scalable framework for routine genomic reanalysis, provided that final interpretation and clinical reporting remain under expert review.

In addition, the present study established and evaluated only the initial implementation of the Talos workflow. We did not assess the likely substantially lower workload of repeated reanalysis cycles. The efficiency gains of iterative automated reanalysis may therefore be greater in practice than reflected here. Reanalysis should therefore not be understood as a fixed one-time event, but as a resource-dependent component of continuous genomic care. From a technical perspective, our study also highlights the need for cleaner and more standardized VCF-processing workflows with greater interoperability between diagnostic pipelines and downstream reanalysis tools.

Beyond workflow efficiency, these findings carry the direct patient-centered implication that periodic automated reanalysis of no-finding and cases with VUS findings should be considered an integral component of rare-disease genomic care. Whole genome sequencing generates a comprehensive dataset that does not lose value when the initial analysis ends. Its diagnostic potential increases as gene–disease associations expand and analytical tools improve. For patients who remain without a molecular diagnosis, this creates a renewed opportunity to resolve the diagnostic odyssey. Therefore, we propose that ongoing reanalysis of these cases should be embedded as standard practice in rare-disease genomic care.

## Abbreviations

ACGS: Association for Clinical Genomic Science
ACMG: American College of Medical Genetics and Genomics
AMP: Association for Molecular Pathology
B/LB: benign/likely benign
CNV: copy-number variant
GS: genome sequencing
HPO: Human Phenotype Ontology
HGVS: Human Genome Variation Society
MOI: mode of inheritance
P/LP: pathogenic/likely pathogenic
SMN: survival motor neuron
SNV: single-nucleotide variant
STR: short tandem repeat
SV: structural variant
UKSH: University Medical Center Schleswig-Holstein
VCF: variant call format
VUS: variant of uncertain significance.

## Supplementary Materials

Supplementary Materials(Preprint).xlsx

Supplementary Table **S1**: Patient Cohort — Clinical and Genomic Overview

Supplementary Table **S2**: Missed by Talos (P/LP)

## Ethics approval and consent to participate

Ethics approval and consent to participate: This study was conducted in full compliance with the ethical standards set forth by the Declaration of Helsinki and was approved by the Ethics Committee of the Institute of Human Genetics, University Hospital Schleswig–Holstein, University of Kiel, Germany (Ethics Approval Number: [D 621/21]). All participants and/or their legal guardians provided written informed consent prior to inclusion in the study. Confidentiality and privacy of all participants were rigorously maintained, with data anonymized to protect personal information. Consent for publication: Not applicable.

## Consent for publication

Not applicable.

## Data availability

Summary-level variant data have been included in the supplementary materials. All clinically relevant variants identified in this study have been submitted to ClinVar [39]. Raw sequencing data are not publicly available due to ethical and legal restrictions but may be available upon request with appropriate approvals. Data can be made available upon reasonable request to the corresponding author. If approved, the data will be provided within a preparation period of up to four weeks.

## Materials availability

Not applicable.

## Code availability

Adapted Talos Code: https://github.com/hexkash/UKSH-TalosSV

## Author contributions

Conceptualization: D.K., M.S.;

Data curation: D.K., V.A., S.M., C.U.T.;

Formal analysis: D.K., V.A., S.M., C.U.T.;

Funding acquisition: M.S.;

Investigation: D.K., V.A., S.M., C.U.T., I.N., I.V., A.C., M.S.;

Methodology: D.K., V.A., S.M., I.N., M.S.;

Project administration: D.K., I.N., M.S.;

Resources: I.N., I.V., A.C., M.S.;

Supervision: I.N., M.S.;

Validation: D.K., V.A., C.U.T., A.C., I.V., I.N., M.S.;

Writing-original draft: D.K.;

Writing-review & editing: D.K., V.A., S.M., C.U.T., A.C., I.V., I.N., M.S.

